# GLP-1 Receptor Agonists and Risk of Paralytic Ileus: A drug-target Mendelian Randomization Study

**DOI:** 10.1101/2024.10.17.24315627

**Authors:** Pingjian Ding, Zhenxiang Gao, Maria P. Gorenflo, Rong Xu

## Abstract

**Background:** Paralytic ileus (PI), a condition characterized by reduced bowel motor activity without physical obstruction, can be affected by complications from type 2 diabetes (T2D) and anti-diabetic medications. It is unclear of the causal associations of glucagon-like peptide-1 receptor agonists (GLP-1RAs) with the risk of PI in the context of T2D management.

**Methods:** To investigate the causal relationship of GLP-1RAs with PI, we conducted a 2-sample mendelian randomization (MR) study based on summary statistics from genome-wide association studies (GWAS). Genetic variants in the GLP1R were identified as genetical proxies of GLP-1RAs by the glycemic control therapy, based on genetic associations with glycated hemoglobin (GWAS n=344,182) and T2D (n_cases/controls_=228,499/1,178,783). The effects of GLP-1RAs were estimated for PI risk (n_cases/controls_=517/182,423) using GWAS data from the FinnGen project.

**Results:** Based on MR analysis, GLP-1RAs are causally associated with a decreased risk of PI (OR per 1 mmol/mol decrease in glycated hemoglobin: 0.21; 95% confidence interval [CI]=0.06-0.69). The magnitude of these benefit exceeded those expected from improved glycemic control more generally.

**Conclusions:** Our study’s findings show that GLP-1RAs are causally associated with a lower risk for PI, which provides information to guide clinicians in the selection of appropriate therapies for individuals with T2D while mitigating the risk of developing PI. Investigating the underlying mechanisms that contribute to the lower PI risk associated with GLP-1RAs is essential for a deeper understanding of these associations.

## 1. Background

Glucagon-like peptide-1 receptor agonists (GLP-1RAs), such as exenatide, liraglutide, dulaglutide, and semaglutide, are medications for managing type 2 diabetes (T2D) [1], [2], [3], [4]. These agents mimic GLP-1, enhancing insulin secretion, suppressing glucagon release, and slowing gastric emptying to improve glycemic control [5], [6], [7]. Individuals with T2D face an elevated risk of gastrointestinal complications, including paralytic ileus (PI) [8], characterized by impaired motor activity in the ileus without physical obstruction [9]. Unlike mechanical obstructions (such as a physical blockage in the intestine), PI affects the entire length of the intestine. Factors related to diabetes, including neuropathy affecting the nerves that control the gastrointestinal tract [10], [11] and changes in gut motility [12], can contribute to the development of PI, making the problem more severe and widespread. Therefore, effective diabetes management, including maintaining optimal glycemic control, is essential to mitigate the risk of complications including PI. GLP-1RAs are highly effective in controlling T2D [1], [2], [3], [4], suggesting their potential beneficial effects on mitigating T2D-associated PI risk.

Observational studies have yielded inconsistent findings of GLP-1RAs on the gastrointestinal system [13], [14]. A recent study [13] suggested that GLP-1RAs, compared to bupropion-naltrexone, were associated with a higher risk of bowel obstruction in participants with obesity. However, when participants with hyperlipidemia were excluded from the analysis, GLP-1RAs were no longer associated with an increased risk of bowel obstruction. A further investigation [14] aimed to estimate whether there is an elevated risk of intestinal obstruction associated with GLP-1RAs and DPP-4 inhibitors compared to sodium-glucose cotransporter-2 (SGLT-2) inhibitors. This study found no association between GLP-1RAs and intestinal obstruction, while DPP-4 inhibitors were associated with an increased risk of obstruction. These inconsistencies may be partially attributable to different comparators used in these observational studies. In addition, no studies have focused on PI. Unlike a mechanical obstruction, PI is caused by a functional impairment in the bowel’s ability to contract, often due to nerve or muscle issues [9]. Among various gastrointestinal events, PI is common in diabetic patients due to this specific nerve dysfunction. Given the limited research on the association between GLP-1RAs and PI, along with the availability of GWAS data for PI, this study conducted a Mendelian randomization (MR) study involving 1,934,404 patients to investigate the potential causal association of GLP-1RAs with PI.

## 2. Methods

We employed a drug-target MR study that includes only variants or near the gene encoding the target of a therapeutic intervention [15], [16] based on GWAS summary statistics to evaluate the effects of genetically proxied GLP-1RAs on PI. The extent to which genetic variations in the GLP-1R gene indicate long-term GLP-1RA responses was determined by examining associations between regional SNPs and hemoglobin A1C (HbA1c) levels as biomarkers of glycemic status [17]. These associations were identified in comprehensive genome-wide association studies (GWAS) of glycated hemoglobin (available from: http://www.nealelab.is/uk-biobank) and type 2 diabetes [18]. Subsequently, associations between selected variants and the PI trait [19], [20] were merged to evaluate the effects of GLP-1RAs on the risk of PI. Figure 1 presents the conceptual MR design.

**Figure 1.**
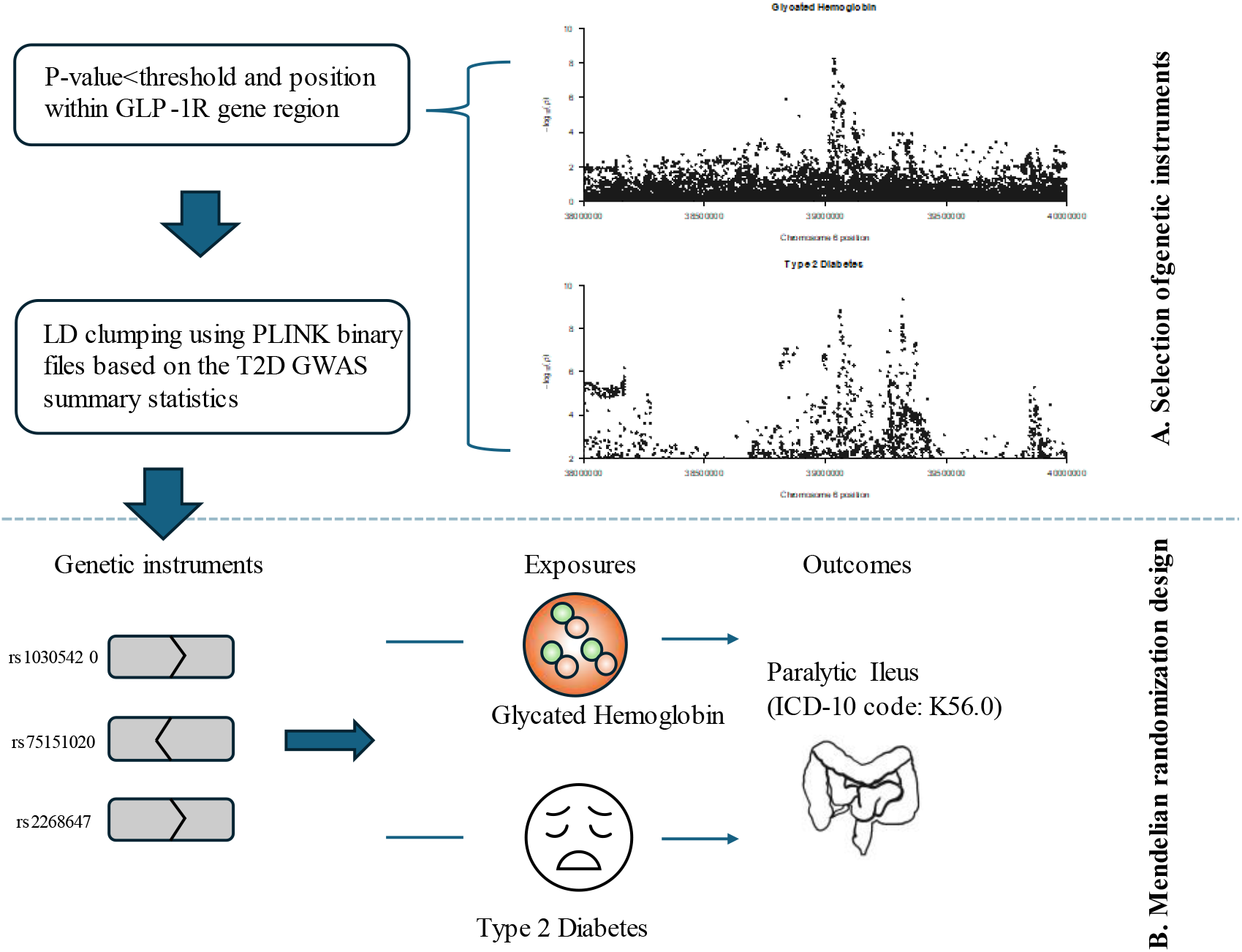
Overview of the MR analysis. A) Selection of genetic instruments. B) Mendelian randomization design.

### 2.1 Genetic proxies for GLP-1RAs and glycemic control

We selected variants associated with T2D at genome-wide significance (P<5*10^−8^) and glycated hemoglobin at nominal significance (P<0.05) that were minimally correlated (linkage disequilibrium threshold of r^2^<0.1 using PLINK) within the GLP-1R gene (build GRCh37: chromosome 6: 39,016,574-39,055,519) [21]. Instrumental variants were selected based on GWAS summary statistics for both T2D and glycated hemoglobin to mitigate the potential bias. The LD clumping method was conducted based on the T2D summary statistics (Figure 1(A)). We obtained genetic associations for T2D from a large meta-analysis comprised of 228,499 patients with T2D and 1,178,783 controls [18]. We used genetic association data for glycated hemoglobin (data field=30750) on UK Biobank (n=344,182) participants (Figure 1(B)). Moreover, the functional consequence was used to annotate these variants, such as intron and missense variants [22]. Tissue-specific gene expression data from Genotype-Tissue Expression (GTEx) v8 [23] was used to determine whether variants were annotated as directionally concordant based on their association with lower glycated hemoglobin and higher expression of GLP-1R (or vice versa). Moreover, we utilized the same association to select genetic variants that were not located within 1 megabase of GLP-1R, which are regarded as genetic proxies for glycemic control using non-GLP-1RA medications. Since many variants were identified for glycemic control using non-GLP-1RA medications, we used a stricter clumping threshold of r2<0.001 and window size=10kb to minimize bias from linkage disequilibrium.

### 2.2. Statistical analysis of the MR study

We obtained genetic association data for PI risk from FinnGen participants with 517 cases and 182,423 controls (Open GWAS ID=finn-b-K11_PARALIL) [19], [20], [24]. The PI case was defined using the ICD-10 code K56.0 (“Paralytic ileus”), while controls were defined as participants without ICD-10 codes K55, K56, K57, K58, K59, K60, K61, K62 or K63. In the MR analysis, we aligned the effect alleles of each instrumental variant to harmonize genetic associations with the exposure and the outcome. We conducted the inverse-variance weighted (IVW) [25] method to calculate the MR estimates for genetically proxied GLP-1RAs and non-GLP-1RA medications. These estimates were oriented towards a reduction in glycated hemoglobin. We conducted all MR analyses using the TwoSampleMR package in R [24]. We performed a comparison to evaluate whether a GLP-1RA could have a better protective effect on PI risk than non-GLP-1RA medications (**eMethods**). We conducted a set of additional analyses to assess the reliability and robustness of the MR findings. First, given that missense variants are frequently presumed to induce disease by affecting protein level and are expected to respond to modulators [26], we constrained the genetic proxies to missense variants specifically within GLP-1R. Second, to assess the sensitivity to our choice on weighting the variants, we also performed additional analyses of weighting the instrumental variants by their associations with T2D instead of glycated hemoglobin [21]. Third, horizontal pleiotropy may bias the MR estimates if the genetic variants proxying GLP-1RAs affect PI through a pathway independent of GLP-1RAs. Thus, we tested for any such potential bias by calculating the Cochran Q test P value [27] to indicate heterogeneity and a MR-Egger regression intercept [25] to assess pleiotropy. We then conducted the single variant tests and leave-one-out analyses to identify high influence points [28].

## 3. Results

### 3.1. GLP-1R variants and their associations with glycated hemoglobin, T2DM, and PI

For the primary analysis, two intron variants (rs2268647 and rs75151020) and a missense variant (rs10305420) were selected to genetically proxy glycated hemoglobin lowering through GLP-1RAs (**Table 1**). The strength of all three instruments, quantified by F-statistics, was greater than 10. Thereinto, through querying through the GTEx v8 database, two of these selected variants, rs10305420 and rs75151020, have significant associations with GLP-1R expression across various human tissues. Notably, both variants were annotated as directionally concordant, as they were linked to lower glycated hemoglobin and higher expression of GLP-1R (or vice versa). The variant rs75151020 has no significant association with gene expression in any of the tissues within the GTEx database. 332 independent variants were available in the PI GWAS summary statistics, serving as genetic proxies for glycemic control using non-GLP-1RA medications.

**Table 1.** Descriptive information on the GLP-1R variants analyzed in the study, and their associations with glycated hemoglobin, T2DM, and PI

|  | rs10305420 | rs75151020 | rs2268647 |
| --- | --- | --- | --- |
| EA/NEA | T/C | C/A | T/C |
| Chr: Position (GRCh37) | 6: 39016636 | 6: 39031592 | 6: 39043178 |
| Functional Consequence | Missense variant | Intron variant | Intron variant |
| Tissues (NES), Concordance | Pancreas (0.24), Yes | N/A | Pancreas (-0.27), Yes |
| Association with traits, beta (SE), P value |  |  |  |
| glycated hemoglobin | -0.051 (0.016), 1.30e-03 | 0.066 (0.015), 1.51e-05 | 0.119 (0.026), 7.08e-06 |
| T2DM | -0.032 (0.004), 5.11e-14 | 0.021 (0.004), 4.95e-08 | 0.041 (0.007), 1.37e-09 |
| PI | -0.139 (0.064), 3.05e-02 | 0.07 (0.063), 2.67e-01 | 0.172 (0.126), 1.71e-01 |
| The instrument strength quantified with R2 and F-statistics |  |  |  |
| glycated hemoglobin | 3.01e-05, 10.34 | 5.86e-05, 20.17 | 5.44e-05, 18.73 |

Abbreviations: GRCh37= Genome Reference Consortium Human Build 37; Chr=Chromosome; EA=Effect Allele; NEA=Non-Effect Allele; NES=Normalized Effect Size; SE=Standard Error; T2DM=Type 2 Diabetes Mellitus; PI= Paralytic Ileus

### 3.2. GLP-1RAs are associated with reduced PI risk

Our MR analysis revealed that genetically proxied GLP-1RAs are associated with a lower risk of PI (odds ratio [OR] per 1 mmol/mol decrease in glycated hemoglobin, 0.205; 95% CI, 0.062-0.685; P=1e-02) (**Figure 2, eFigure 1**). Given that missense variants are commonly presumed to induce disease by affecting the protein level and are expected to respond to modulators [26], we conducted separated analyses to investigate the impact of genetically predicted GLP-1RAs on PI risk. The MR estimate remained consistent in the MR analysis when only utilizing the missense variant rs10305420 (OR, 0.066 95% CI, 0.006-0.774; P=3.05e-02). Excluding this missense variant and relying solely on intron variants, the MR analysis yielded statistically insignificant results (OR, 0.294 95% CI, 0.074-1.167; P=8.17e-02). This insignificance may be attributed to the limited power of the two intron variants to detect the effect. Single variant analyses and leave-one-out analyses suggest that the MR estimate is not solely derived by a single variant (**eFigure 2-3**). On the other hand, our MR analysis suggests no evidence for an association between genetically proxied non-GLP-1RA medications and PI risk (OR, 1.047 95% CI, 0.974-1.124; P=2.13e-01). This OR for genetically proxied non-GLP-1RA medications was significantly larger than that of genetically proxied GLP-1RAs (P_difference_=8.17e-03).

We weighted the genetic proxies for GLP-1RAs by T2D liability instead of glycated hemoglobin, and the analysis suggested similar evidence for a reduction in PI risk (OR per log-odds decrease in T2D liability, 0.017; 95% CI, 0.001, 0.303; P=5.54e-03) (**eFigure 4-5**). Additionally, similar findings were observed with weighting by T2D when only missense variants or intron variants considered (**eFigure 4, 6, 7**). Cochran Q test and MR-Egger regression intercept suggest that these findings of weighting by either glycated hemoglobin or T2D liability were not due to heterogeneity (Cochran Q P>0.05) or horizontal pleiotropy (MR-Egger intercept P>0.05) (**Figure 2 and eFigure 4**).

**Figure 2.**
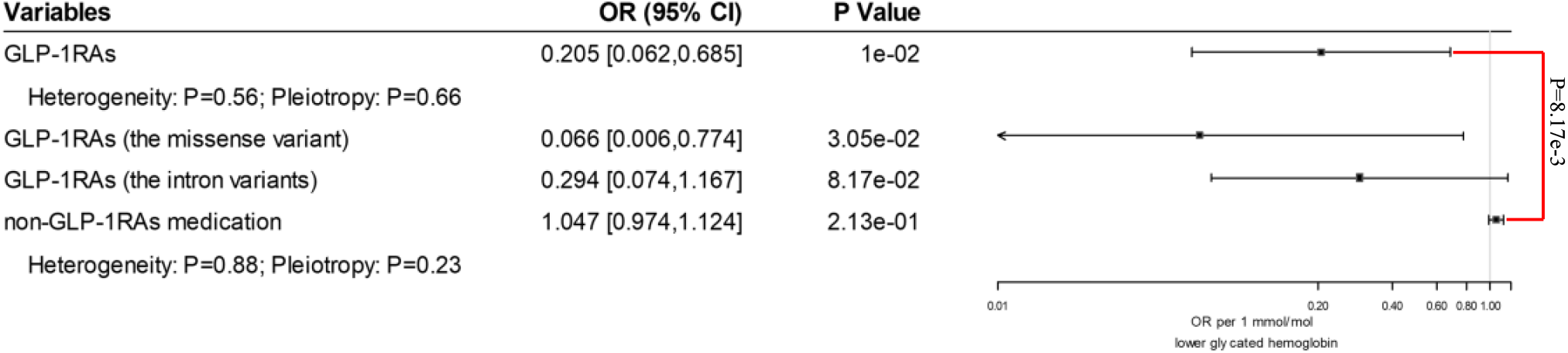
Forest plot depicting Mendelian randomization estimates for the association of genetically proxied GLP-1RAs and non-GLP-1RA medications with risk of PI. Estimates reflect the effect of a reduction in glycated hemoglobin on PI risk.

## 4. Discussion

Utilizing summary-level data from more than 1.9 million individuals and conducting multiple sensitivity analyses, this comprehensive study demonstrates a causal association between GLP-1RAs and a reduced risk of PI. To the best of our knowledge, this is the first large-scale Mendelian randomization (MR) analysis to explore potential causal links between GLP-1RAs and PI risk. In a previous cohort study comparing patients with obesity treated with GLP-1RAs and those with bupropion-naltrexone, Sodhi et al. [13] found that GLP-1RAs were associated with an increased risk of bowel obstruction and the association diminished after excluding patients with hyperlipidemia. Faillie et al. [14] showed that GLP-1RAs were associated with an increased risk of intestinal obstruction, compared to SGLT-2 inhibitors. Distinct from bowel obstruction that involves a mechanical or functional blockage in the intestines which prevents the normal movement of digested products [29], PI is a functional issue affecting the muscles and nerves, mimicking the symptoms of intestinal obstruction even in the absence of any physical blockage [9], [30]. Our study focused on PI, primarily due to the limited prior research exploring the association between GLP-1RAs and PI, as well as the availability of GWAS data for PI. Our study extends previous epidemiological studies showing that GLP-1RAs are causally associated with reduced PI risk compared with non-GLP-1RA anti-diabetic medications.

The observed protective association between GLP1-RAs and PI in the context of glycated hemoglobin lowering may be due to the efficacy of GLP-1RAs in lowering blood glucose, which could mitigate hyperglycemic mesenteric microvascular damage and therefore improve blood flow to the ileum [31], reducing the risk for PI. Strategies for managing T2DM often include maintaining optimal blood glucose levels, which can help reduce the risk of inflammation [32] and neuropathy [10]. The relationship between GLP-1RAs and the potential decrease in PI risk may involve several additional mechanisms. Various studies have indicated that GLP-1RAs have anti-inflammatory properties, potentially influencing the immune system [33].Disruption of regular gastrointestinal motility can be caused by inflammation [34]. GLP-1RAs, through their anti-inflammatory effects, may play a role in decreasing the risk of PI. GLP-1RAs have been implicated in neuroprotection [35], although effects on the enteric nervous system have not been explored. Neuroprotection might help prevent damage to the nerves controlling intestinal function, reducing the risk of PI.

In this study, we employed drug target MR analysis showing that GLP-1RAs may have therapeutic benefits in reducing PI risk through lowering glycated hemoglobin. Drug target MR allows researchers to infer causality between a genetic variant linked to drug target and disease outcomes. It also offers insights into mechanisms of action by determining whether altering a specific gene can influence disease risk. Drug target MR studies have been employed for drug discovery such as PCSK9 inhibition for cognitive function [36], IL-6 inhibition for Alzheimer’s disease [37], and ACE inhibitors for mental disorders [38]. However, there are limitations in drug target MR studies, including the limited availability of GWAS studies for specific drug targets. In addition, our approach is constrained by the limited number of genetic proxies available to GLP-1RAs, which would preclude further sensitivity analyses for horizontal pleiotropy [39]. Since proper glycemic control helps diabetes-related complications, this MR study primarily focuses on GLP-1RA’s effect on PI through its glycemic effects. However, GLP-1RAs exhibit diverse effects beyond glycemic control, including promoting weight loss and reducing inflammation. Further research is needed to investigate the potential causal links between GLP-1RAs and PI through other underlying mechanisms.

## 5. Conclusion

Our mendelian randomization studies provided genetic evidence to support that GLP-1RAs are associated with reduced risk of developing PI. Our findings provide information to guide clinicians in the selection of appropriate therapies for individuals with T2DM while mitigating the risk of developing PI. Investigating the underlying mechanisms that contribute to the lower PI risk associated with GLP-1RAs is essential for a deeper understanding of these associations.

## Supporting information

Supplemental Online Content

## Data Availability

All data produced in the present work are contained in the manuscript.

## Abbreviation

PI: Paralytic ileus
T2D: Type 2 diabetes
GLP-1RAs: Glucagon-like peptide-1 receptor agonists
MR: Mendelian randomization
GWAS: Genome-wide association studies
SGLT-2: Sodium-glucose cotransporter-2
HbA1c: hemoglobin A1C
GTEx: Genotype-Tissue Expression
IVW: Inverse-variance weighted
GRCh37: Genome Reference Consortium Human Build 37
Chr: Chromosome
EA: Effect Allele
NEA: Non-Effect Allele
NES: Normalized Effect Size
SE: Standard Error
OR: Odds ratio
LD: Linkage Disequilibrium.

## Declarations

The authors declare that they have no conflict of interest.

## Ethics approval and consent to participate

Not applicable.

## Consent for publication

Not applicable.

## Author’s contributions

PD: conceptualization, methodology, data curation, conducted analysis, writing—original draft, writing—review and editing and visualization. ZG: supported interpretations, writing—original draft, writing—review and editing and visualization. MG: supported interpretations and writing—review and editing. RX: conceptualization, supervision, supported interpretations, funding acquisition and writing—review and editing. All authors read and approved the final manuscript.

## Data Availability

The original contributions presented in the study are included in the article/supplementary material.

## Acknowledgements

The authors would like to thank the researchers for providing the valuable summarized data.

