## Supplemental Online Content for "GLP-1 Receptor Agonists and Risk of Paralytic Ileus: A drug-target Mendelian Randomization Study"

**eMethods.**

**eFigure 1.** Scatter plot displaying genetic associations of the GLP1R genetic proxies with glycated hemoglobin (mmol/mol, x-axis) and paralytic ileus (log-odds, y-axis).

**eFigure 2.** The plot shows the effect as estimated using each of the variants on their own, and comparing against the effect as estimated using the IVW method that use all the SNPs.

**eFigure 3.** Leave-one-out sensitivity analysis of MR estimates between the GLP1R genetic proxies with glycated hemoglobin and paralytic ileus.

**eFigure 4.** Forest plot depicting the MR estimates reflecting the effect of a reduction in T2DM on PI risk.

**eFigure 5.** Scatter plot displaying genetic associations of the GLP1R genetic proxies with type 2 diabetes mellitus liability (log-odds, x-axis) and paralytic ileus (log-odds, y-axis).

**eFigure 6.** The plot shows the effect as estimated using each of the variants on their own, and comparing against the effect as estimated using the IVW method that use all the SNPs.

**eFigure 7.** Leave-one-out sensitivity analysis of MR estimates between the GLP1R genetic proxies with type 2 diabetes mellitus and paralytic ileus.

**eMethods.**

1. A comparison to evaluate whether a GLP-1RAs could have a better protective effect on PI risk than non-GLP-1RAs medication

Under the null hypothesis of equality of two MR beta coefficients following a standard unit normal, we evaluated the significance of the difference between the MR beta coefficient $\beta_{GLP-1RAs}$ and the MR beta coefficient $\beta_{non-GLP-1RAs}$ as follows [26]:

$z=\left( \beta_{GLP-1RAs}-\beta_{non-GLP-1RAs} \right)/\left[ {SE}^{2}\left( \beta_{GLP-1RAs} \right)+{SE}^{2}\left( \beta_{non-GLP-1RAs} \right) \right]^{1/2}$,

where the estimate for this difference (numerator) was obtained by taking the difference between two MR estimates, and the standard error of the difference (denominator) is the square root of the sum of the two squared standard errors. All hypothesis tests were 2 sided.

**eFigure 1**. Scatter plot displaying genetic associations of the GLP1R genetic proxies with glycated hemoglobin (mmol/mol, x-axis) and paralytic ileus (log-odds, y-axis).

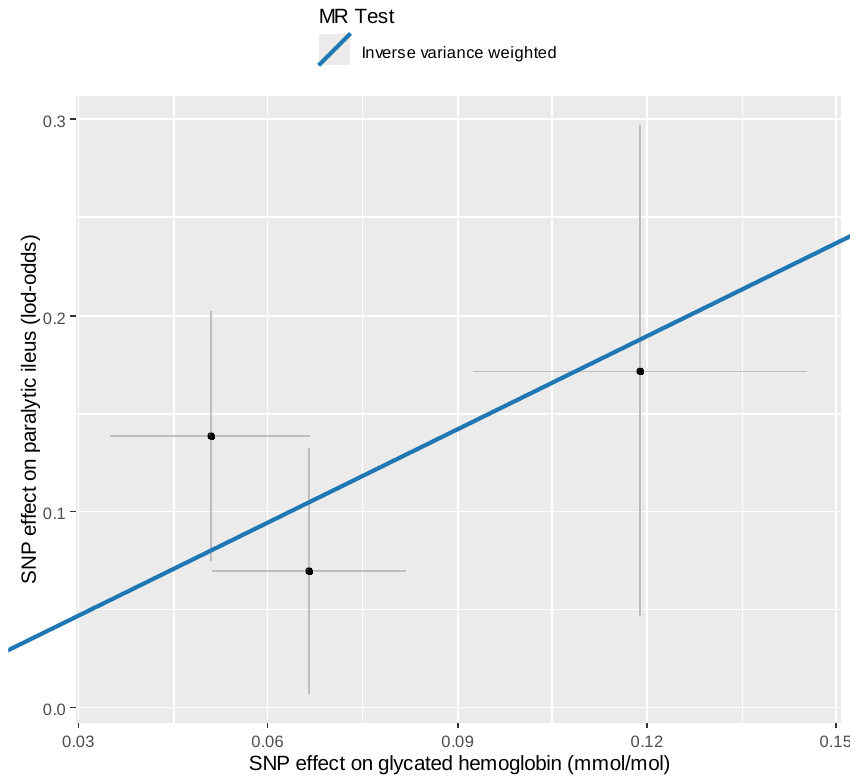

**eFigure 2**. The plot shows the effect as estimated using each of the variants on their own, and comparing against the effect as estimated using the IVW method that use all the SNPs.

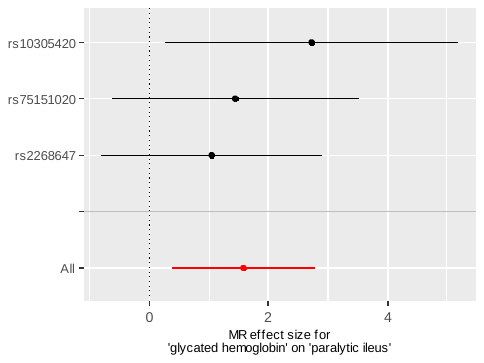

**eFigure 3**. Leave-one-out sensitivity analysis of MR estimates between the GLP1R genetic proxies with glycated hemoglobin and paralytic ileus.

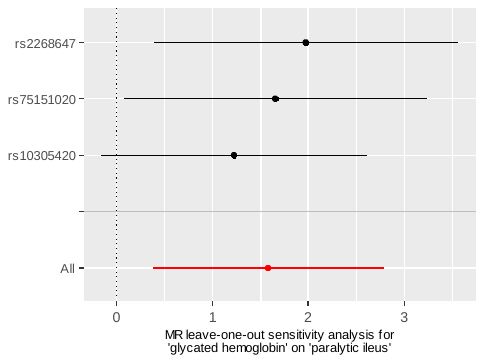

**eFigure 4**. Forest plot depicting the MR estimates reflecting the effect of a reduction in T2DM on PI risk.

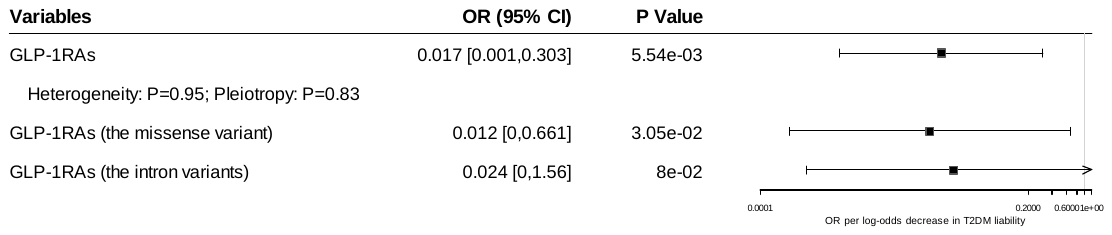

**eFigure 5**. Scatter plot displaying genetic associations of the GLP1R genetic proxies with type 2 diabetes mellitus liability (log-odds, x-axis) and paralytic ileus (log-odds, y-axis).

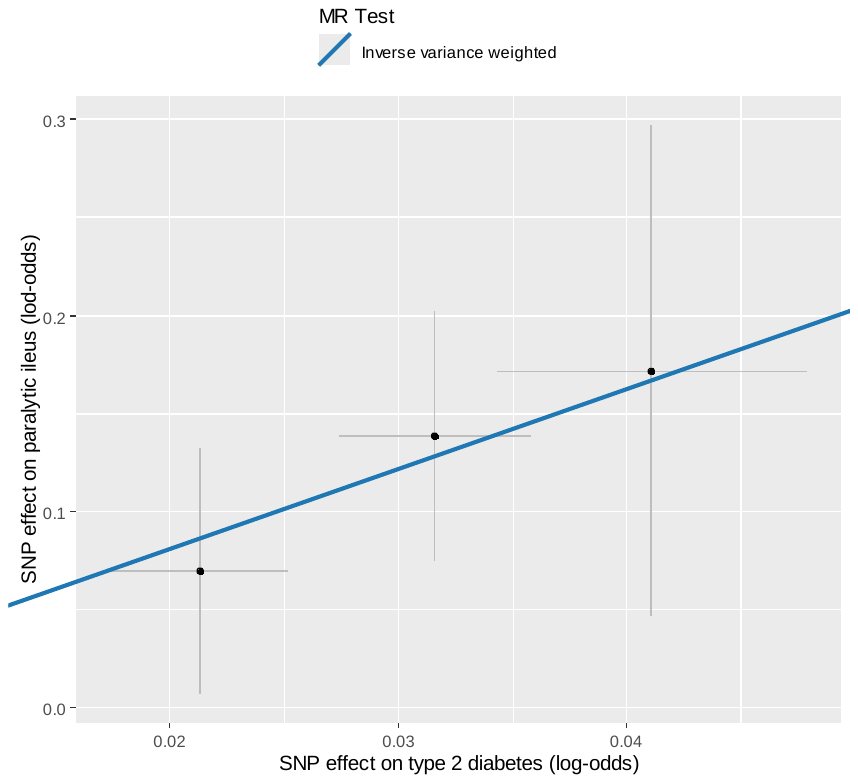

**eFigure 6**. The plot shows the effect as estimated using each of the variants on their own, and comparing against the effect as estimated using the IVW method that use all the SNPs.

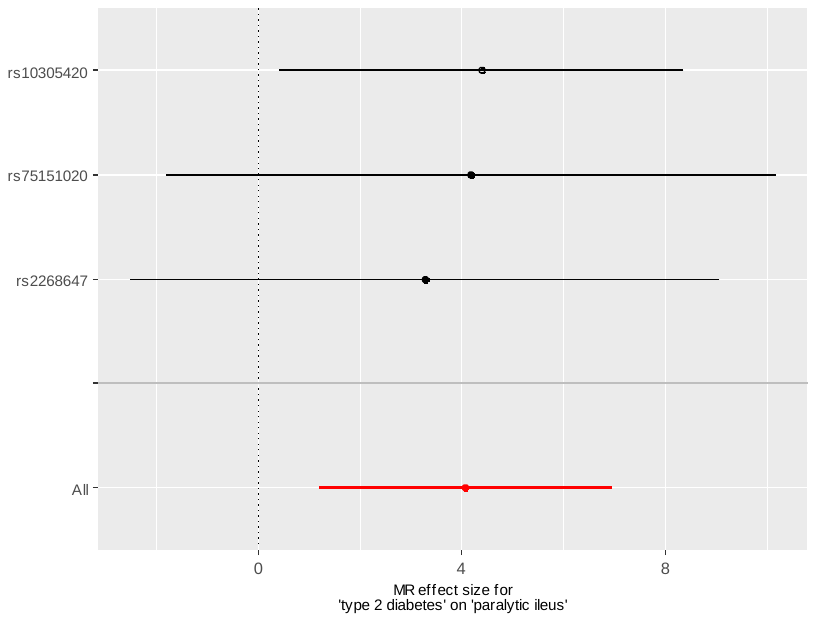

**eFigure 7**. Leave-one-out sensitivity analysis of MR estimates between the GLP1R genetic proxies with type 2 diabetes mellitus and paralytic ileus.

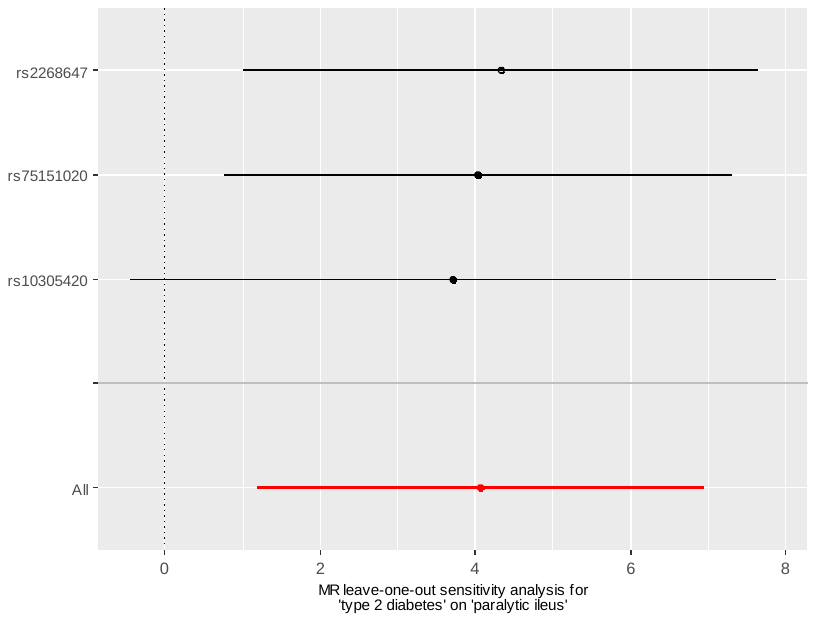
